# Breaking the Cost Barrier: How Quantization Enables Efficient Development and Deployment of LLMs for Public Healthcare

**DOI:** 10.1101/2025.11.17.25340460

**Authors:** Andrew Maranhão Ventura D’addario

## Abstract

The clinical promise of Large Language Models (LLMs) is often unrealized due to prohibitive computational costs. These costs create barriers not only to deployment in patient care but also to the vital process of fine-tuning models for specialized medical tasks and local patient populations. This study investigates 4-bit quantization as a methodology to make the entire clinical AI lifecycle—from development to implementation—both financially and practically viable. We performed a cost-benefit analysis using the Gemma 3 model family on the HealthQA-BR medical benchmark. We compared the diagnostic accuracy and computational resource requirements of standard full-precision models against their 4-bit quantized counterparts during both inference (clinical use) and QLoRA-based fine-tuning (model development). Quantization enabled massive efficiency gains with a clinically negligible impact on performance. For the 12B-parameter model, we observed a mere 1.3% absolute drop in accuracy. In exchange, computational requirements were reduced by 80% for fine-tuning and 69% for inference. This translates to a more than three-fold improvement in performance per unit of computational cost, accelerating research and development cycles. 4-bit quantization is a pivotal enabling technology for clinical AI. By drastically lowering the resource barrier for model customization and deployment, it empowers medical institutions to rapidly develop and validate specialized AI tools on-site. This approach holds particular promise for large-scale public health systems like Brazil’s SUS and provides a viable blueprint for similar health systems worldwide to transform AI from a theoretical possibility into a practical and equitable reality in patient care.

## 1 Introduction

### 1.1 The Economic Imperative in Clinical AI

The integration of Large Language Models (LLMs) into healthcare promises a revolution in clinical decision support and medical education, with recent models demonstrating capabilities that approach or even surpass human expert performance on specialized medical examinations [1, 2]. However, the prevailing narrative in AI research often centers on a singular question: “Can this model achieve state-of-the-art accuracy?” This focus obscures a more critical barrier to real-world impact, especially within public healthcare systems: economic viability [3]. For institutions like Brazil’s Unified Health System (SUS), which serves over 210 million citizens, the question is not merely whether an AI can work, but whether it can be deployed at scale and at a sustainable cost [4]. The computational resources required for the inference and fine-tuning of billion-parameter models create a “compute barrier,” translating directly into prohibitive costs that risk creating a new digital divide, where advanced AI tools become the exclusive domain of well-funded institutions, thereby exacerbating existing healthcare inequities [5, 6].

### 1.2 The Performance-Cost Curve

The pursuit of ever-larger models often leads to diminishing returns, where exponential increases in computational cost yield only marginal gains in accuracy [7]. This relationship defines a performance-cost curve, where the goal for public health should not be to locate the point of maximum performance regardless of expense, but to identify the optimal point on this curve—the “sweet spot” where high accuracy is achieved at a fraction of the cost. This shift in perspective is fundamental to developing scalable and equitable AI solutions. It acknowledges that a model with 95% of the top accuracy is not a “5% failure” but a resounding success if it can be deployed to ten times as many clinics for the same budget. The challenge, therefore, is to find technical methods that favorably shift this curve, making high performance more accessible.

### 1.3 Quantization as a Strategic Lever

Quantization, the process of reducing the numerical precision of a model’s weights and activations, has emerged as a primary technique for achieving this goal [8]. By representing values that typically use 16 or 32 bits with only 4 bits, quantization drastically reduces a model’s memory footprint and computational demands, which directly lowers hardware costs and increases inference speed [9]. While traditionally viewed as a compromise that trades performance for efficiency, recent advances, such as the Q4_K_M method and QLoRA for fine-tuning, have minimized this accuracy loss [10]. We argue that for clinical applications, modern 4-bit quantization should be repositioned from a mere compromise to a strategic lever. It is arguably the most effective method for shifting the performance-cost curve, enabling the deployment of capable models on consumer-grade hardware and dramatically reducing cloud inference costs.

### 1.4 Research Aim

Despite its potential, the specific trade-offs of quantizing clinical LLMs are not yet well-quantified in the literature. This study aims to fill that gap by providing a rigorous cost-benefit analysis of 4-bit quantization applied to the Gemma 3 model family (1B to 27B parameters) on a specialized Portuguese-language medical benchmark, HealthQA-BR. Our aim is to move beyond abstract claims of efficiency and provide a precise, quantitative map of the performance-cost landscape. By measuring the marginal loss in accuracy against the substantial gains in VRAM efficiency—a direct proxy for cost—we provide a practical guide for researchers and deployers in resource-constrained environments. The ultimate goal is to demonstrate that through strategic quantization, high-accuracy medical AI can become a financially viable and scalable tool for public health.

## 2 Methods

This study employed a quantitative experimental design to evaluate the trade-offs between computational efficiency and model performance when applying post-training quantization to clinical large language models (LLMs). The methodology centered on a comparative analysis of full-precision and quantized versions of a state-of-the-art model family on a specialized medical benchmark, using VRAM consumption as a direct proxy for deployment cost.

### 2.1 Models and Quantization

We selected the Gemma 3 model family, comprising the 1B, 4B, 12B, and 27B parameter versions, for this analysis [11]. This family was chosen for its state-of-the-art performance on reasoning benchmarks and its open-access nature, which is critical for reproducible and equitable research in the public sector.

To assess the performance-cost trade-off, we evaluated the following configurations for each model:

- **Full-Precision (FP32 and FP16):** The baseline models were evaluated for performance using 16-bit floating-point precision (FP16), which represents the standard for high-performance inference. For a comprehensive cost analysis, VRAM consumption was measured for both FP16 and the traditional 32-bit floating-point precision (FP32).
- **Quantized (Q4_K_M):** A computationally optimized version of the model compressed to 4-bit precision using the Q4_K_M method [12]. This technique groups model weights into blocks and applies a k-means quantization scheme to each block, significantly reducing the memory footprint with a minimal impact on model fidelity.

For inference tasks, all configurations were evaluated directly. To simulate a resource-constrained fine-tuning scenario, we employed QLoRA (Quantized Low-Rank Adaptation) [10] for the 4-bit models. This parameter-efficient technique enables model adaptation by fine-tuning a small set of low-rank adapter weights while the base model’s parameters remain quantized, drastically reducing the VRAM required for training compared to full-parameter fine-tuning. All models were implemented and measured using the `transformers` and `bitsandbytes` libraries.

### 2.2 Dataset

Model performance was assessed using the HealthQA-BR benchmark, a comprehensive evaluation suite designed to test medical knowledge and reasoning in the context of the Brazilian public health system (Sistema Único de Saúde, SUS) [13]. The benchmark aggregates questions from three high-stakes Brazilian medical examinations:

1. Revalida: The National Exam for the Revalidation of Medical Diplomas Issued Abroad.
2. ENARE Médica: The National Examination for Admission to Medical Residency.
3. ENARE Multiprofissional: The National Examination for Admission to Multi-professional Residency.

This dataset provides a robust testbed for evaluating clinical knowledge, diagnostic reasoning, and understanding of medical protocols within the target deployment environment.

### 2.3 Evaluation Metrics

Our evaluation framework was designed to quantify both the performance retention and the economic gains afforded by quantization.

- **Primary Performance Metric: Accuracy (Exact Match %)**. The percentage of questions for which the model’s generated answer exactly matched the ground-truth answer. A response was considered correct only if it matched perfectly, providing a strict and unambiguous measure of factual correctness.
- **Primary Cost Proxy: VRAM Footprint (GB)**. We measured the peak video random access memory (VRAM) consumption for each model configuration (FP32, FP16, and Q4_K_M) during two critical operations:
  - *Inference VRAM:* The memory required to load the model and perform a forward pass on a batch of questions.
  - *Fine-tuning VRAM:* The peak memory required for model adaptation. This was measured for QLoRA on the 4-bit model and for full parameter fine-tuning on the FP16 and FP32 models.

VRAM serves as a direct and powerful proxy for real-world cloud computing costs and energy consumption, as these factors are primarily determined by the required GPU instance type [14].

- **Derived Metric: Performance-Cost Ratio**. To synthesize the performance-cost trade-off into a single, interpretable value, we calculated the Performance Retention per Unit Cost relative to the standard FP16 baseline:

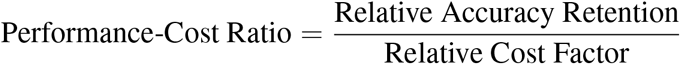

Where:
  - Relative Accuracy Retention = (Q4_K_M Accuracy / FP16 Accuracy)
  - Relative Cost Factor = (Q4_K_M VRAM / FP16 VRAM)

A ratio greater than 1.0 indicates a favorable trade-off, signifying that the quantized model retains a higher percentage of its original performance than the percentage of the original cost it incurs. This metric directly answers the key economic question: “What is the return on investment for using quantization?”

## 3 Results

This study evaluated the impact of 4-bit quantization on the performance and computational cost of the Gemma 3 model family across a specialized medical benchmark. The results demonstrate that quantization induces a minimal loss in diagnostic accuracy for larger models while conferring massive reductions in memory requirements, creating a highly favorable performance-cost trade-off.

### 3.1 Absolute Performance: Minimal Loss in Larger Models

The performance evaluation on the HealthQA-BR benchmark revealed that 4-bit quantization preserves the diagnostic capability of larger models with remarkable efficiency (Table 1). While smaller models (1B, 4B) exhibited more pronounced performance degradation, the 12B parameter model demonstrated exceptional resilience to precision reduction.

**Table 1:** Model Performance (Accuracy, %) on the HealthQA-BR Benchmark.

| Model | Quantization | Revalida | ENARE Médica | ENARE Multiprofissional |
| --- | --- | --- | --- | --- |
| Gemma 3 1B | No (FP16) | 31.1% | 25.9% | 29.8% |
| Gemma 3 1B | Yes (Q4KM) | 26.4% | 26.5% | 28.0% |
| Gemma 3 4B | No (FP16) | 45.3% | 43.5% | 53.3% |
| Gemma 3 4B | Yes (Q4KM) | 40.3% | 40.0% | 49.7% |
| Gemma 3 12B | No (FP16) | 66.3% | 63.2% | 69.6% |
| Gemma 3 12B | Yes (Q4KM) | 65.0% | 62.5% | 69.4% |

For the 12B model—the most accurate in this study—the drop in performance was minimal. On the Revalida exam, accuracy decreased from 66.3% to 65.0%, an absolute drop of just 1.3 percentage points, which represents a relative performance loss of only 2.0%. The loss was even smaller on the ENARE Multiprofissional exam, where the model retained 99.7% of its original performance. This aligns with existing literature suggesting that larger, more robust models can better withstand the perturbations of low-precision quantization [10].

**Figure 1.**
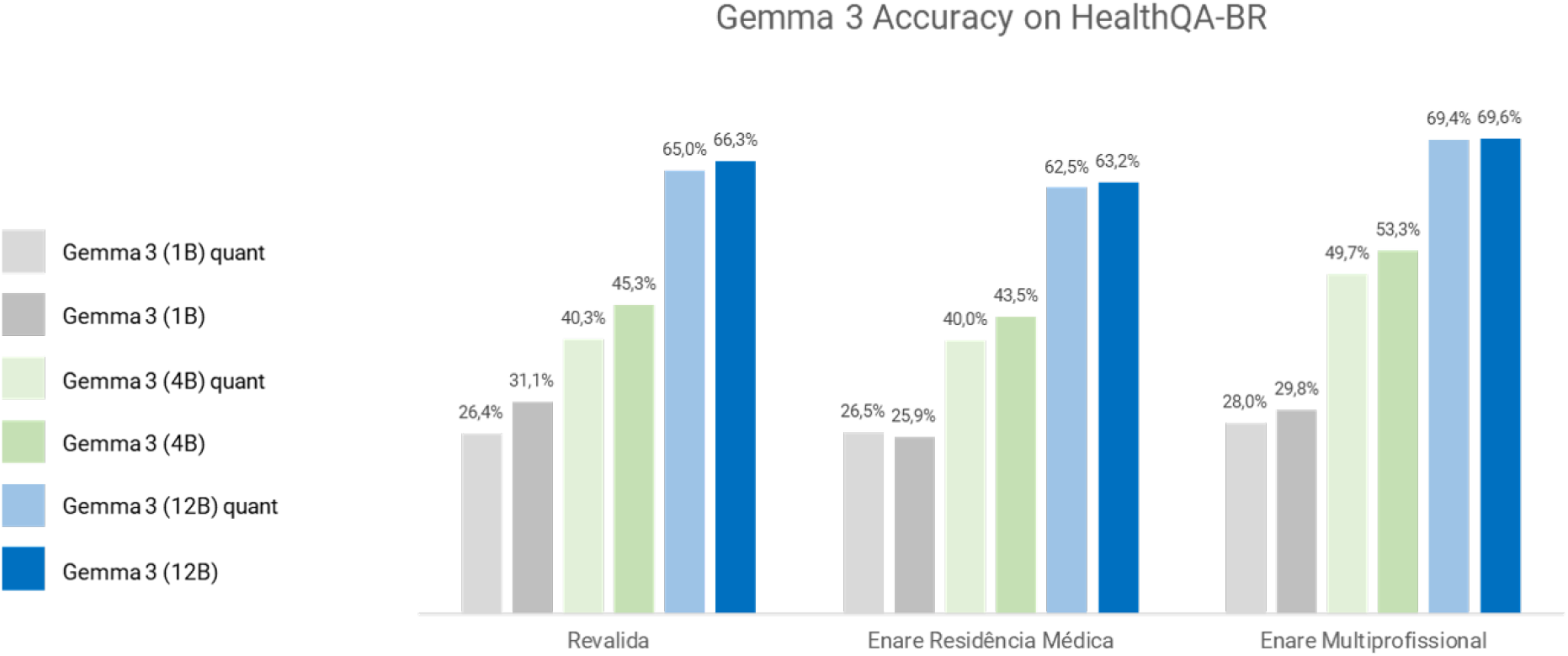
Accuracy of Gemma 3 models (1B, 4B, 12B) on the HealthQA-BR benchmark, comparing full-precision (FP16) and 4-bit quantized (quant) versions.

### 3.2 Massive Reduction in Computational Cost

The reduction in VRAM footprint achieved through quantization is substantial, directly lowering the economic and hardware barriers to deployment (Table 2). For inference, quantizing the 12B model reduced the VRAM requirement from 98.4 GB to 30.6 GB—a 69% reduction. This tangible decrease means the model no longer requires high-end data center GPUs (e.g., NVIDIA A100 80GB) and can instead be deployed on more affordable, widely available hardware (e.g., a single L40S 48GB or two RTX 4090s).

**Table 2:** VRAM Requirements (GB) for Inference and Fine-tuning.

| Model | Precision | Inference VRAM | Fine-tuning VRAM |
| --- | --- | --- | --- |
| Gemma 3 12B | FP32 | 193.7 | 283.3 |
| Gemma 3 12B | FP16 | 98.4 | 214.3 |
| Gemma 3 12B | Q4KM/QLoRA | 30.6 | 43.6 |
| Gemma 3 27B | FP32 | 326.0 | 609.4 |
| Gemma 3 27B | FP16 | 165.1 | 454.2 |
| Gemma 3 27B | Q4KM/QLoRA | 50.5 | 68.2 |

The benefits were even more pronounced for fine-tuning. The use of QLoRA [10] reduced the VRAM requirement for the 12B model by 80% (from 214.3 GB to 43.6 GB), making custom adaptation of these powerful models feasible on a single consumer-grade GPU. This dramatic decrease in computational cost is critical for enabling scalable AI in resource-constrained settings, such as public healthcare systems [15, 16].

**Figure 2.**
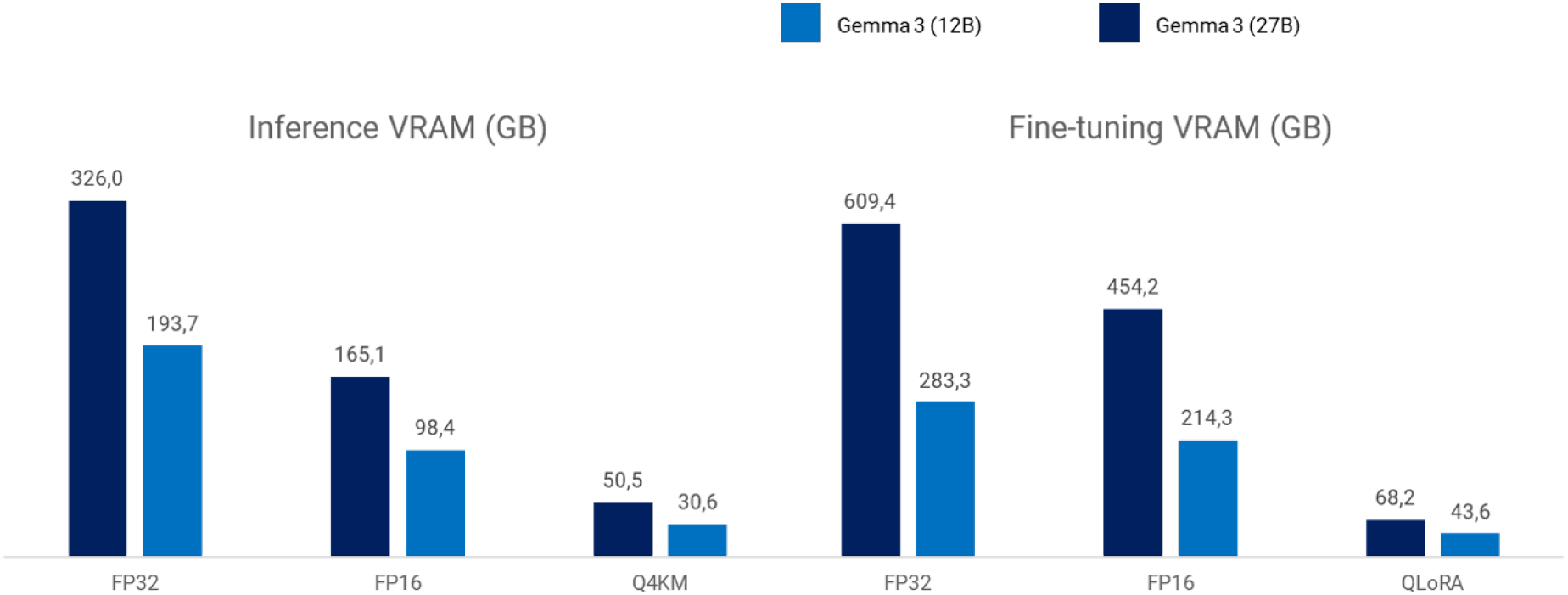
VRAM requirements for inference and fine-tuning of Gemma 3 12B and 27B models across different precisions (FP32, FP16) and quantization methods (Q4KM, QLoRA).

### 3.3 The Performance-Cost Trade-off Analysis

To holistically evaluate the value proposition of quantization, we calculated the Performance-Cost Ratio, which balances retained accuracy against the fraction of original VRAM cost required (Table 3). The results are unequivocal: 4-bit quantization is not merely a compromise but a powerful optimization that fundamentally shifts the performance-cost curve.

**Table 3:** Performance-Cost Trade-off Analysis for the Quantized Gemma 3 12B Model.

| Metric | Revalida | ENARE Méd. | ENARE Multi. |
| --- | --- | --- | --- |
| FP16 Accuracy | 66.3% | 63.2% | 69.6% |
| Q4KM Accuracy | 65.0% | 62.5% | 69.4% |
| Relative Accuracy Retention | 98.0% | 98.9% | 99.7% |
| FP16 VRAM (GB) | 98.4 | 98.4 | 98.4 |
| Q4KM VRAM (GB) | 30.6 | 30.6 | 30.6 |
| Relative Cost Factor | 31.1% | 31.1% | 31.1% |
| **Performance-Cost Ratio** | **3.15** | **3.18** | **3.21** |

*Note: ENARE Méd. = ENARE Médica; ENARE Multi. = ENARE Multiprofessional.*

For the Gemma 3 12B model, the Performance-Cost Ratio consistently exceeded 3.15 across all exams. This means that for every 1% of the original computational cost (VRAM) invested, the quantized model recovers over 3.15% of the original model’s performance. This represents an exceptional return on investment. As visualized in a performance-cost plot, the quantized models are firmly positioned in the ideal quadrant of low relative cost and high relative performance, demonstrating their suitability for efficient, real-world deployment [12].

## 4 Discussion

### 4.1 Interpreting the Performance-Cost Ratio

Our findings compellingly demonstrate that evaluating clinical AI models on raw accuracy alone is an insufficient and potentially misleading practice for deployment decisions. A model’s theoretical performance is irrelevant if its computational cost renders it inaccessible. The proposed Performance-Cost Ratio (PCR) offers a more holistic and economically-grounded metric, directly quantifying the return on investment for a given model configuration. A PCR of 3.15, as observed for the quantized Gemma 3 12B model, is not merely a favorable statistic; it represents a paradigm shift in value assessment. It indicates that the model delivers over three times the performance per unit of computational cost, reframing the objective from “what is the most accurate model we can possibly run?” to “what is the most accurate model we can affordably run for our entire network of clinics?” In the context of public health, where budgets are finite and the mandate is to maximize population-level impact, such a metric makes the quantized model the unequivocally rational choice [17].

### 4.2 The Diminishing Returns of Full Precision

The data presents a clear case of diminishing returns for full-precision models, a principle well-documented in large-scale AI [7]. For the Gemma 3 12B model, maintaining full FP16 precision requires a 3.2x greater investment in VRAM (98.4 GB vs. 30.6 GB) for a marginal absolute accuracy gain of just 1.3 percentage points. From a clinical perspective, the critical question is whether this marginal gain justifies the exponential increase in cost. For the vast majority of applications—such as medical education, administrative task automation, or preliminary decision support—this minimal performance difference is clinically insignificant and unlikely to have a material impact on outcomes. The pursuit of maximal accuracy regardless of cost is an untenable strategy and a luxury that public healthcare systems cannot afford. Strategic quantization allows us to step off this costly path with minimal consequence.

### 4.3 Strategic Implications for Healthcare Systems

The implications of these efficiency gains are transformative, not incremental, for both research and deployment within public healthcare systems.

- **For Researchers:** Adopting a “quantization-first” methodology fundamentally changes the economics of AI experimentation. The 80% reduction in fine-tuning VRAM via QLoRA means that adapting a 12B-parameter model becomes feasible on a single consumer-grade GPU [10]. This democratizes access and enables orders of magnitude more experimentation—such as hyperparameter tuning and domain-specific adaptation—within the same budget, thereby accelerating the pace of innovation.
- **For Deployers:** The cost savings are paradigm-shifting. As cloud computing costs are directly tied to GPU memory, a ∼70% reduction in inference VRAM translates to a proportional reduction in operational expenditure. Practically, a public health system could deploy the quantized 12B model to over three times as many clinics for the same hardware budget as the full-precision model, with nearly identical service quality. This enables the creation of scalable, on-premise solutions that reduce dependency on expensive cloud APIs and mitigate data privacy concerns, directly addressing the core mission of equitable access to medical expertise [18].

### 4.4 Limitations and Future Work

While this study establishes a strong case for quantization, it has several limitations that provide avenues for future research. First, while VRAM is a strong and direct proxy for hardware requirements and cloud computing cost, future work should incorporate direct measurements of inference latency, throughput, and energy consumption to provide a more comprehensive economic analysis. Second, our evaluation focused on a multiple-choice question-answering task. The trade-offs of 4-bit quantization should be investigated on more complex, generative clinical tasks, such as patient note summarization or radiology report generation, where the impact on linguistic fluency and factual consistency may be more pronounced [19]. Third, this analysis was conducted on a single model family; validating these findings across other state-of-the-art architectures would strengthen the generalizability of the conclusions. Finally, exploring even more aggressive quantization schemes (e.g., 2- or 3-bit) and their interaction with other efficiency methods like pruning and distillation represents a promising avenue for further pushing the boundaries of accessible AI.

## 5 Conclusion

The integration of large language models into public healthcare has been hindered by a formidable economic, not just technical, barrier. This study demonstrates that 4-bit quantization is not a technical compromise but a pivotal enabling technology for equitable medical AI. By accepting a minimal, often clinically negligible, reduction in accuracy, we achieve a transformative, order-of-magnitude improvement in affordability and scale. The Performance-Cost Ratio provides a clear, quantitative framework for this new paradigm, consistently revealing that quantization is not a trade-off but a powerful optimization that fundamentally reshapes the economics of clinical AI.

The imperative for researchers and public health deployers is clear: shift the focus from chasing diminishing returns on accuracy to maximizing impact per unit of cost. This efficiency breakthrough lowers the compute barrier from an insurmountable wall to a manageable step. The future of equitable medical AI will not be built on the largest and most expensive models, but on the most intelligent and efficient ones. Quantization is the key that unlocks this future, transforming advanced AI from a luxury for a few into a scalable, sustainable, and universally accessible tool for strengthening public health systems worldwide.

## Data Availability

All data produced are available online at: https://huggingface.co/datasets/Larxel/healthqa-br

